# Timing of Complete Multivessel Revascularization in Patients Presenting with Non-ST-Elevation Acute Coronary Syndrome

**DOI:** 10.1101/2023.05.29.23290502

**Authors:** Jacob J. Elscot, Hala Kakar, Paola Scarparo, Wijnand K. den Dekker, Johan Bennett, Carl E. Schotborgh, Rene van der Schaaf, Manel Sabaté, Raúl Moreno, Koen Ameloot, Rutger J. van Bommel, Daniele Forlani, Bert Van Reet, Giovanni Esposito, Maurits T. Dirksen, Willem P.T. Ruifrok, Bert R. C. Everaert, Carlos Van Mieghem, Eduardo Pinar, Fernando Alfonso, Paul Cummins, Mattie Lenzen, Salvatore Brugaletta, Joost Daemen, Eric Boersma, Nicolas M. Van Mieghem, Roberto Diletti, BIOVASC Investigators

## Abstract

**Background:** Multivessel coronary artery disease (MVD) is highly prevalent in patients presenting with non-ST-segment elevation myocardial infarction (NSTE-ACS) and is associated with worse clinical outcomes compared with single vessel disease patients. Complete revascularization of the culprit and all significant non-culprit lesions reduces the incidence of major adverse cardiac events, but the optimal timing of non-culprit artery revascularization remains unclear.

**Methods:** This prespecified substudy of the randomized BIOVASC trial included patients who presented with NSTE-ACS and MVD, defined as ≥ 1 non-culprit related coronary artery with a diameter of ≥ 2.5 mm and ≥ 70% stenosis as per visual estimation or positive coronary physiology testing. Risk differences of the composite of all-cause mortality, myocardial infarction, unplanned ischemia driven revascularization or cerebrovascular events and its individual components were compared between the patients who were randomized to immediate and staged complete revascularization at 30 days and 1 year.

**Results:** The BIOVASC trial enrolled 1525 patients, 917 patients presented with NSTE-ACS, of whom 459 were allocated to the immediate complete and 458 to the staged complete revascularization group. The incidences of the primary composite outcome were similar in the two groups (7.9% vs. 10.1%, risk difference 2.2%, 95%CI −1.5 to 6.0, p = 0.24). Immediate complete revascularization was associated with a significant reduction in the incidence of myocardial infarction (2.0% vs. 5.3%, risk difference 3.3%, 95% confidence interval [CI] 0.9 to 5.7, p = 0.008), which was maintained after exclusion of procedure related myocardial infarctions occurring at the index or staged procedure (2.0% vs. 4.4%, risk difference 2.4%, 95%CI 0.1 to 4.7, p = 0.039). Unplanned ischemia driven revascularizations were also reduced in the immediate complete revascularization group (4.2% vs. 7.8%, risk difference 3.5%, 95%CI 0.4 to 6.6, p = 0.025).

**Conclusions:** Immediate complete revascularization is safe in patients with NSTE-ACS and MVD and was associated with a reduction in myocardial infarctions and unplanned ischemia driven revascularizations at 1 year.

**Clinical Perspective:** *What Is New?*

- This prespecified subanalysis of the BIOVASC trial shows that all spontaneous myocardial infarctions between the index and staged procedure occurred in the population of patients that initially presented with NSTE-ACS. At 30 days and 1 year patients randomized to immediate complete revascularization have fewer myocardial infarctions and unplanned ischemia driven revascularizations.

*What Are the Clinical Implications?*

- Immediate complete revascularization appears to be a safe strategy and can be a reasonable option for complete revascularization in patients presenting with NSTE-ACS and multivessel disease
- In patients presenting with NSTE-ACS and multivessel disease, misjudgment of the culprit lesion or presence of multiple vulnerable plaques could have a role in the reduction of early occurring myocardial infarctions when performing an immediate complete strategy.

## Introduction

Multivessel coronary artery disease (MVD) is common in patients presenting with an acute coronary syndrome (ACS) without persistent ST-elevations (NSTE-ACS). About 50% of the patients present with one or more significant non-culprit lesions, a condition associated with a higher risk of myocardial infarction (MI), repeat revascularization and mortality^1–5^. An early invasive strategy is beneficial over a conservative approach in terms of better clinical outcomes, especially in high risk patients^6–10^. Several retrospective studies suggested that complete revascularization of both culprit and non-culprit lesions is associated with lower cumulative mortality rates and risk of major adverse cardiac events^3, 11–13^. Therefore, recent guidelines report that complete revascularization should be considered in patients with MVD and NSTE-ACS, tailored to patients’ characteristics, preferences and comorbidities^14^. However, the ideal timing of non-culprit revascularization in an immediate or staged setting remains unclear. The ESC guidelines provide a class IIb recommendation for complete revascularization during index percutaneous coronary intervention (PCI) ^14^ based on one small randomized trial showing a lower risk of MACE, driven by a lower repeat revascularization rate when immediate complete revascularization (ICR) was performed instead of staged complete revascularization (SCR) ^15^.

The recently published BIOVASC randomized trial showed that ICR is non-inferior to SCR in terms of a composite of all-cause mortality, MI, any unplanned ischemia-driven revascularization or cerebrovascular events in patients presenting with ACS at 1 year post index procedure. ^16^

Against this background, we now present the trial results in the subcohort of NSTE-ACS patients, which was prespecified in the protocol.

## Methods

### Protocol Design and Randomization

The BIOVASC trial was a multicenter, investigator-initiated, open-label randomized controlled non-inferiority trial with participating sites in the Netherlands, Belgium, Italy and Spain, comparing ICR with SCR in patients presenting with ACS and MVD. Details of the trial design and the main results have been previously reported^16, 17^. In summary, 1525 patients presenting with acute coronary syndrome including both ST segment elevation myocardial infarction (STEMI) and NSTE-ACS and multivessel MVD, defined as at least 70% stenosis in a non-culprit vessel ≥ 2.5 mm in diameter by visual estimation or positive coronary physiology testing, were randomized in a 1:1 ratio to ICR or SCR within 6 weeks after index procedure. Invasive coronary imaging or physiology assessment was performed at the operator’s discretion. Exclusion criteria consisted of the absence of a clear culprit, previous coronary artery bypass grafting, cardiogenic shock and the presence of a chronic total occlusion in a vessel ≥ 2.5 mm in diameter. The primary endpoint was a composite of all-cause mortality, nonfatal myocardial infarction, any unplanned ischemia-driven revascularization, and cerebrovascular events at 1 year post index procedure.

**Figure 1.**
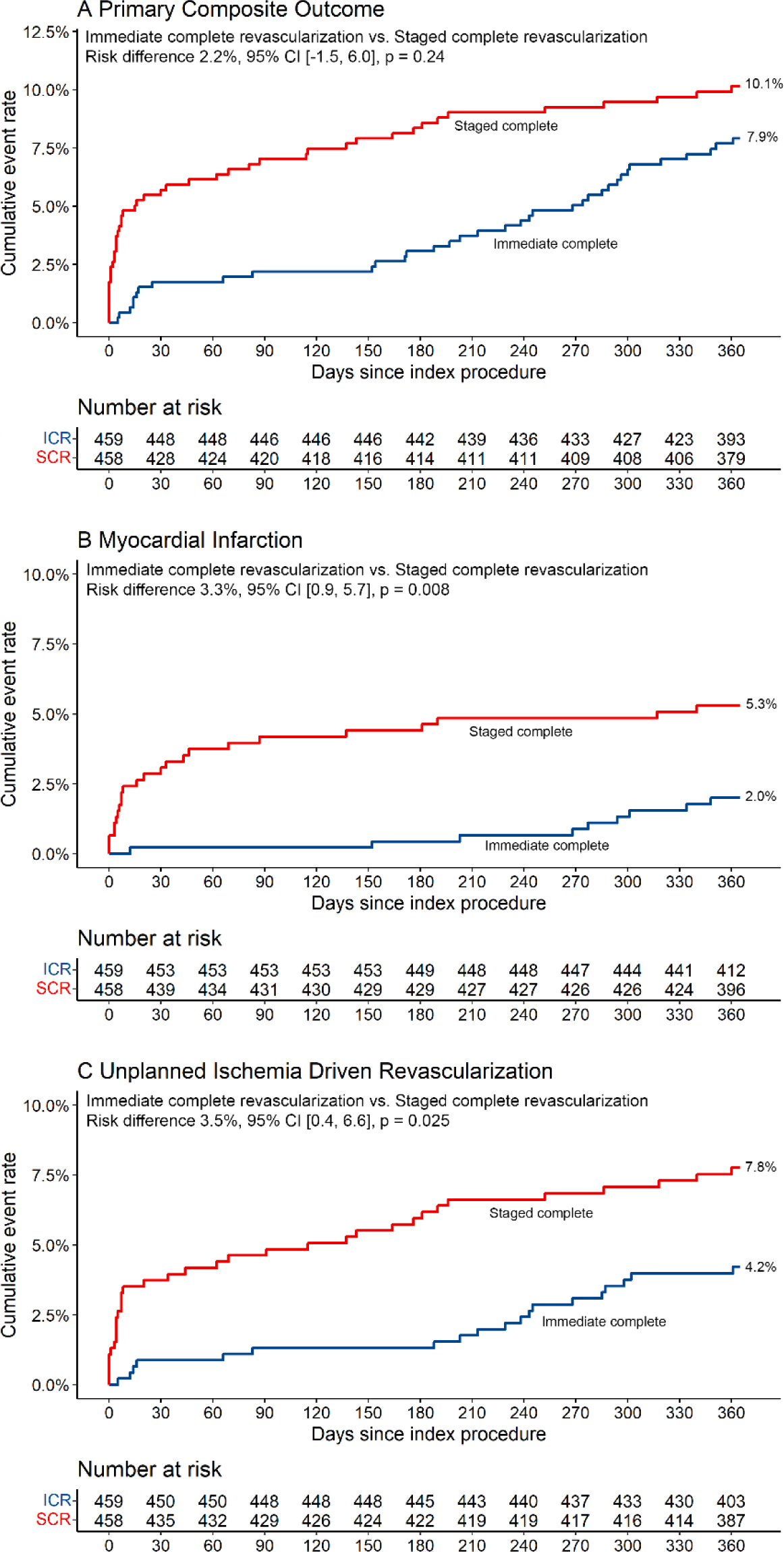
Outcomes The primary outcome is a composite of all-cause mortality, myocardial infarction, unplanned ischemia driven revascularization and cerebrovascular events. A difference in favour of immediate complete revascularization is presented as a positive value. ICR indicates immediate complete revascularization; SCR, staged complete revascularization.

### Prespecified analysis in patients with NSTE-ACS

This BIOVASC substudy is a prespecified analysis designed to ascertain if there was a difference in clinical outcomes when comparing ICR with SCR in the NSTE-ACS population. NSTE-ACS was defined according to current guidelines^14^. In brief, a patient was considered presenting with NSTE-ACS if at least two of the following criteria were present: 1) History consistent with new, or worsening ischemia, occurring at rest or with minimal activity; 2) Coronary angiography with indication to PCI; 3) Electrocardiographic changes compatible with ischemia but not diagnostic for ST-segment elevation myocardial infarction, (i.e. ST depression of 1 mm or greater in two contiguous leads, T-wave inversion more than 3 mm, or any dynamic ST shifts). If cardiomyocyte necrosis was present or absent, a patient would be categorized as presenting with non-ST-segment elevation myocardial infarction (NSTEMI) or unstable angina (UA), respectively.

### Study endpoints

Definitions of all efficacy and safety outcomes have been previously published in detail^17^. Deaths were classified as cardiovascular or non-cardiovascular. If the cause of death was undetermined, it was considered cardiovascular. The definition of MI was in line with the Third Universal Definition^18^, including a modification taking into account the ACS setting similarly to the COMPLETE trial^19^. Repeat revascularization had to be considered both unplanned and ischemia driven to be counted as an endpoint. A clinical events committee, comprising three independent physicians with expertise in interventional cardiology or neurology, adjudicated all potential endpoints.

The primary outcome of the current analysis was a composite all-cause mortality, MI, unplanned ischemia driven revascularization and cerebrovascular events, similar to the main trial. Secondary outcomes include the individual components of the primary outcome composite and a composite of cardiovascular death and myocardial infarction.

**Figure 2.**
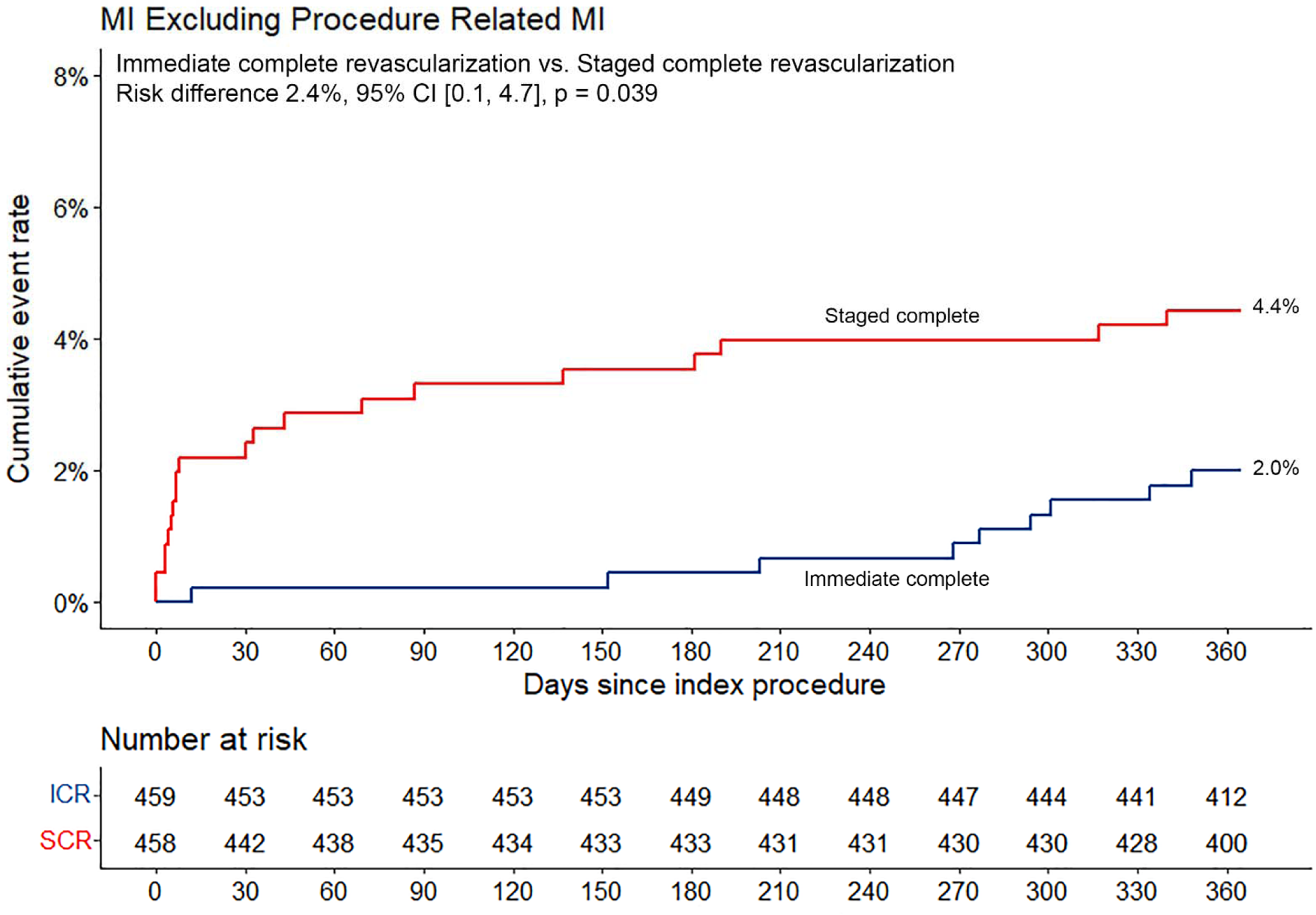
Myocardial Infarction Excluding Procedure Related Myocardial Infarctions Type 4a myocardial infarctions related to the index and staged procedure were excluded from the analysis. A difference in favour of immediate complete revascularization is presented as a positive value. ICR indicates immediate complete revascularization; SCR, staged complete revascularization.

### Statistical Analysis

All randomized patients presenting with NSTE-ACS were included in the analysis as per an intention-to-treat principle. Categorical data were presented as counts and percentages and tested by the chi-square test or Fisher exact test if there was an expected cell value < 5. Continuous data were presented as mean and standard deviation if a Gaussian distribution was present and tested by the unpaired t-test. Alternatively, continuous data were presented as median and quartiles [Q1, Q3] and compared using the Mann-Whitney U test. The distribution of continuous data was tested with the use of the Shapiro-Wilk test.

Cumulative time-to-event curves were calculated with the use of the Kaplan-Meier method. Patients were censored after the first event had occurred or, if event-free, at the date on which they were last known to be alive. Cox proportional hazard regression (PH) was conducted to further explore the relation between randomly allocated treatment and study endpoints. Hazard ratios (HR) were presented with 95% confidence intervals and calculated with use of Cox regression analyses. Assessment of the log-minus log survival plot led to a suspicion of a violated PH assumption for the primary endpoint. Further testing of the Schoenfeld residuals concluded that the PH assumption was not met. Therefore P values for all endpoints were computed on the difference in the cumulative incidence between the two groups for consistency. A two-sided P value < 0.05 was considered statistically significant. All analyses were performed using R version 4.2.1 (packages used: data.table, dplyr, ggplot2, ggpubr, graphics, lubridate, stats, survival, survminer, tidycmprsk).

**Figure 3.**
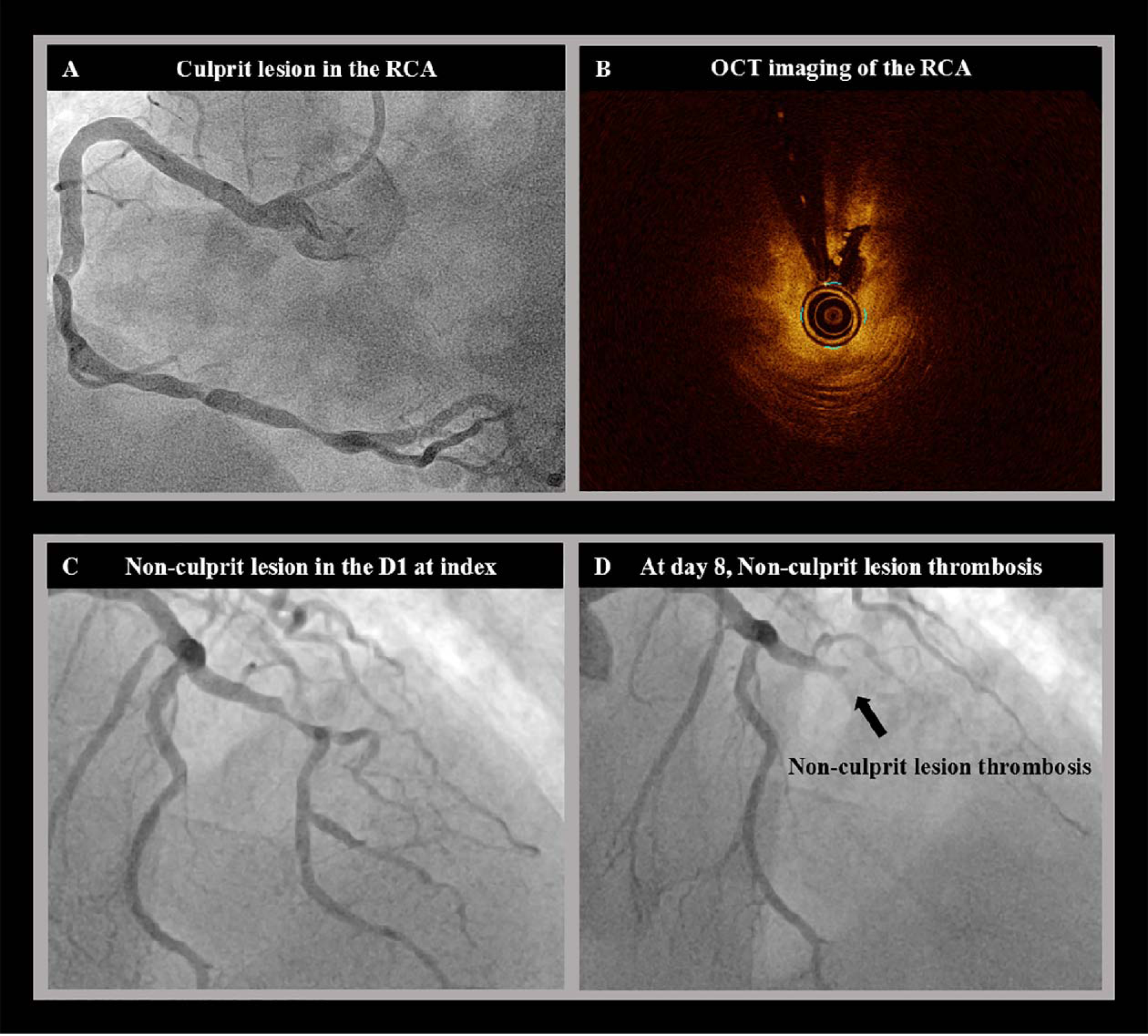
Thrombosis of Non-Culprit in Between the Index and Staged Procedure. A patient in their 70s presented with a NSTEMI. Coronary angiogram revealed subtotal lesions in the RCA (panel A), a significant lesion in the diagonal branch (panel C) and in-stent restenosis of the LCX. After intravascular imaging the lesion in the RCA was identified as the culprit lesion (panel B). The patient was randomized to staged complete revascularization. The RCA was treated successfully and non-culprit lesion treatment was planned after 14 days. At day 8 post index PCI, the patient presented at the emergency due to chest pain. New coronary angiography showed that the significant non-culprit lesion in the diagonal had evolved into a thrombotic occlusion (panel D). D1 indicates first diagonal; LCX, left circumflex artery; NSTEMI, non-ST-elevation myocardial infarction; OCT, optical coherence tomography; PCI, percutaneous coronary intervention; RCA, right coronary artery.

## Results

### Patient characteristics

The BIOVASC trial enrolled 1525 patients, of whom 917 (60.1%) presented with a NSTEMI or UA, with 459 and 458 patients randomized to ICR and and SCR, respectively. ICR and SCR showed similar baseline characteristics (Table 1). Investigator reported complete revascularization was more prevalent in the patients randomized to ICR, despite intracoronary physiology and imaging being more frequently used in those randomized to SCR (Table 2). Additionally, ICR was associated with a lower total stent length, contrast use, radiation dose and a shorter in-hospital stay.

**Table 1.** Baseline Characteristics.

| Characteristics | Immediate Complete<br>Revascularization<br>(N=459) | Staged Complete<br>Revascularization<br>(N=458) | P Value |
| --- | --- | --- | --- |
| Age, years | 67.0 (58.1–74.3) | 66.8 (59.3–73.9) | 0.62 |
| Male sex | 350 (76.3%) | 355 (77.5%) | 0.65 |
| BMI | 27.3 (24.5–30.4) | 27.5 (25.0–30.0) | 0.80 |
| Presentation |  |  | 0.25 |
| NSTEMI | 402 (87.6%) | 388 (84.7%) |  |
| UA | 57 (12.4%) | 70 (15.3%) |  |
| Medical history |  |  |  |
| Previous PCI | 61 (13.3%) | 82 (17.9%) | 0.054 |
| History of MI | 53/458 (11.6%) | 65/458 (14.2%) | 0.24 |
| Peripheral artery disease | 27 (5.9%) | 23 (5.0%) | 0.57 |
| COPD | 38 (8.3%) | 34 (7.4%) | 0.63 |
| Atrial fibrillation or<br>flutter | 23 (5.0%) | 17 (3.7%) | 0.34 |
| Renal insufficiency | 32 (7.0%) | 31 (6.8%) | 0.90 |
| History of stroke | 25 (5.5%) | 18 (3.9%) | 0.28 |
| Hypertension | 286 (62.3%) | 266 (58.1%) | 0.19 |
| Diabetes | 107 (23.3%) | 117 (25.5%) | 0.43 |
| Hypercholesterolemia | 261/457 (57.1%) | 270 (59.0%) | 0.57 |
| Family history of CVD | 150/458 (32.8%) | 151/451 (33.5%) | 0.82 |
| Smoking behavior |  |  | 0.57 |
| Never | 216/455 (47.5%) | 218/454 (48.0%) |  |
| Current | 144/455 (31.6%) | 131/454 (28.9%) |  |
| Former | 95/455 (20.9%) | 105/454 (23.1%) |  |

Data are presented as median (Q1, Q3), n (%), or n/N (%). BMI indicates body-mass index; COPD, chronic obstructive coronary disease; CVD, cardiovascular disease; IQR, interquartile range; MI, myocardial infarction; NSTEMI, non-ST-segment elevation myocardial infarction; PCI, percutaneous coronary intervention; UA, unstable angina

**Table 2.**
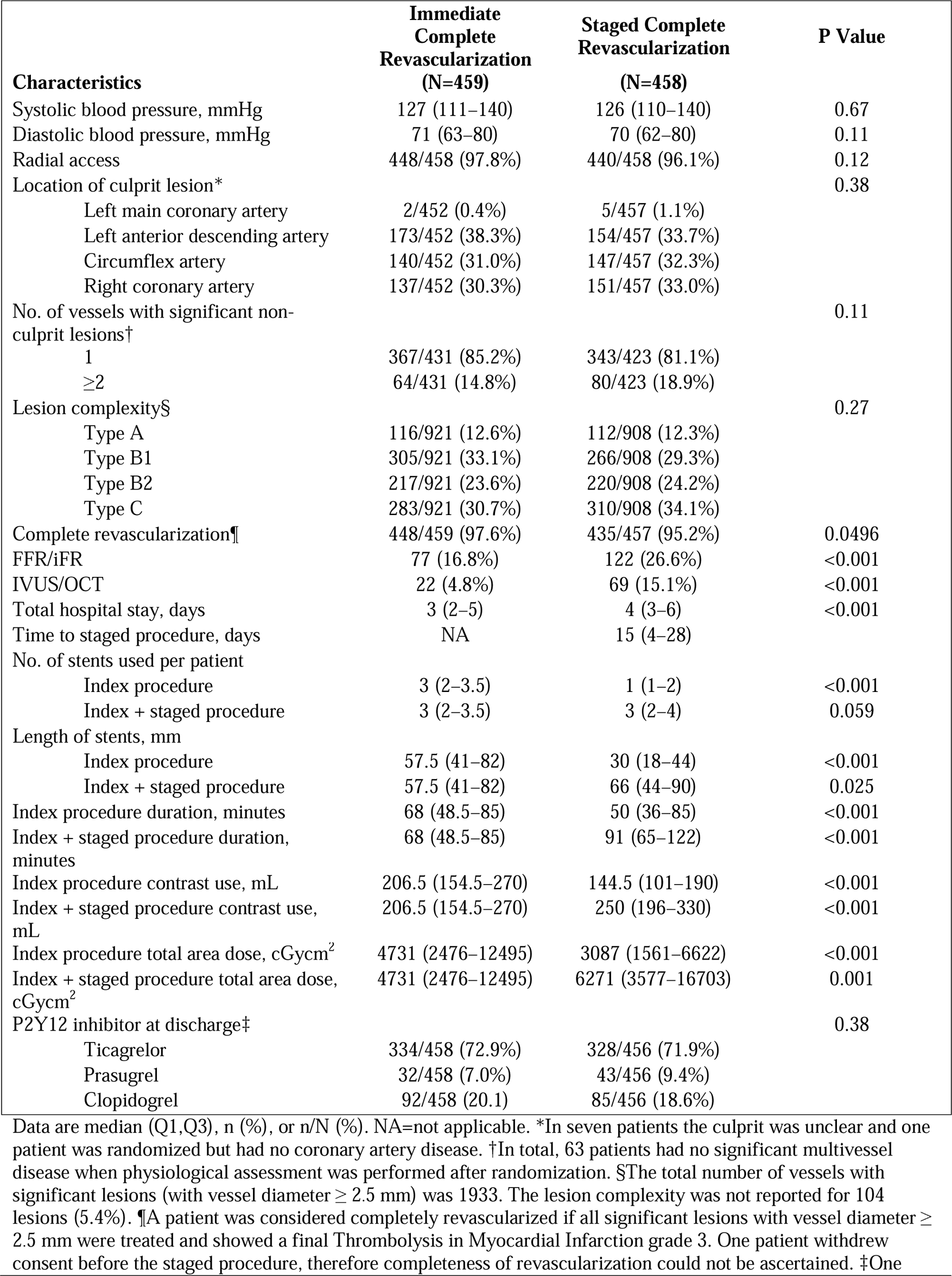
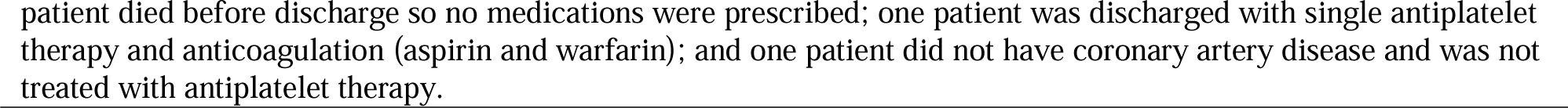
Procedural Characteristics

| **Characteristics** | **Immediate Complete Revascularization (N=459)** | **Staged Complete Revascularization (N=458)** | **P Value** |
| --- | --- | --- | --- |
| Systolic blood pressure, mmHg | 127 (111–140) | 126 (110–140) | 0.67 |
| Diastolic blood pressure, mmHg | 71 (63–80) | 70 (62–80) | 0.11 |
| Radial access | 448/458 (97.8%) | 440/458 (96.1%) | 0.12 |
| Location of culprit lesion* |  |  | 0.38 |
| Left main coronary artery | 2/452 (0.4%) | 5/457 (1.1%) |  |
| Left anterior descending artery | 173/452 (38.3%) | 154/457 (33.7%) |  |
| Circumflex artery | 140/452 (31.0%) | 147/457 (32.3%) |  |
| Right coronary artery | 137/452 (30.3%) | 151/457 (33.0%) |  |
| No. of vessels with significant non-culprit lesions† |  |  | 0.11 |
| 1 | 367/431 (85.2%) | 343/423 (81.1%) |  |
| ≥2 | 64/431 (14.8%) | 80/423 (18.9%) |  |
| Lesion complexity§ |  |  | 0.27 |
| Type A | 116/921 (12.6%) | 112/908 (12.3%) |  |
| Type B1 | 305/921 (33.1%) | 266/908 (29.3%) |  |
| Type B2 | 217/921 (23.6%) | 220/908 (24.2%) |  |
| Type C | 283/921 (30.7%) | 310/908 (34.1%) |  |
| Complete revascularization¶ | 448/459 (97.6%) | 435/457 (95.2%) | 0.0496 |
| FFR/iFR | 77 (16.8%) | 122 (26.6%) | <0.001 |
| IVUS/OCT | 22 (4.8%) | 69 (15.1%) | <0.001 |
| Total hospital stay, days | 3 (2–5) | 4 (3–6) | <0.001 |
| Time to staged procedure, days | NA | 15 (4–28) |  |
| No. of stents used per patient |  |  |  |
| Index procedure | 3 (2–3.5) | 1 (1–2) | <0.001 |
| Index + staged procedure | 3 (2–3.5) | 3 (2–4) | 0.059 |
| Length of stents, mm |  |  |  |
| Index procedure | 57.5 (41–82) | 30 (18–44) | <0.001 |
| Index + staged procedure | 57.5 (41–82) | 66 (44–90) | 0.025 |
| Index procedure duration, minutes | 68 (48.5–85) | 50 (36–85) | <0.001 |
| Index + staged procedure duration, minutes | 68 (48.5–85) | 91 (65–122) | <0.001 |
| Index procedure contrast use, mL | 206.5 (154.5–270) | 144.5 (101–190) | <0.001 |
| Index + staged procedure contrast use, mL | 206.5 (154.5–270) | 250 (196–330) | <0.001 |
| Index procedure total area dose, cGycm<sup>2</sup> | 4731 (2476–12495) | 3087 (1561–6622) | <0.001 |
| Index + staged procedure total area dose, cGycm<sup>2</sup> | 4731 (2476–12495) | 6271 (3577–16703) | 0.001 |
| P2Y12 inhibitor at discharge‡ |  |  | 0.38 |
| Ticagrelor | 334/458 (72.9%) | 328/456 (71.9%) |  |
| Prasugrel | 32/458 (7.0%) | 43/456 (9.4%) |  |
| Clopidogrel | 92/458 (20.1) | 85/456 (18.6%) |  |

Data are median (Q1,Q3), n (%), or n/N (%). NA=not applicable. \*In seven patients the culprit was unclear and one patient was randomized but had no coronary artery disease. †In total, 63 patients had no significant multivessel disease when physiological assessment was performed after randomization. §The total number of vessels with significant lesions (with vessel diameter ≥ 2.5 mm) was 1933. The lesion complexity was not reported for 104 lesions (5.4%). ¶A patient was considered completely revascularized if all significant lesions with vessel diameter ≥ 2.5 mm were treated and showed a final Thrombolysis in Myocardial Infarction grade 3. One patient withdrew consent before the staged procedure, therefore completeness of revascularization could not be ascertained. ‡One
patient died before discharge so no medications were prescribed; one patient was discharged with single antiplatelet therapy and anticoagulation (aspirin and warfarin); and one patient did not have coronary artery disease and was not treated with antiplatelet therapy.

### Outcomes

Follow up was complete in 456 (99.3%) and 452 (98.6%) patients randomized to ICR and SCR respectively.

At 30 days post index procedure, the primary composite outcome (1.8% vs. 5.7%, risk difference 4.0%, 95% confidence interval [CI] 1.5 to 6.4, p = 0.002) and the composite of cardiovascular death and MI (0.2% vs. 3.1%, risk difference 2.9%, 95%CI 1.1 to 4.6, p = 0.001) showed a statistically significant difference in favor of the patients randomized to ICR. The incidence of MI (0.2% vs. 3.1%, risk difference 2.9%, 95%CI 1.2 to 4.5, p < 0.001) and unplanned ischemia driven revascularization (0.9% vs. 3.7%, risk difference 2.9%, 95%CI 0.9 to 4.8, p = 0.004) was also lower in the patients randomized to ICR at 30-day follow-up. All spontaneous MIs between the index and staged procedure occurred in patients that initially presented with NSTE-ACS. Additionally, there was a higher incidence of the composite of all-cause mortality, MI, stroke or major bleeding (BARC 3 or 5) in the SCR arm (1.3% vs. 5.7%, risk difference 4.4%, 95%CI 2.0 to 6.8, p < 0.001). The primary and secondary outcomes at 30 days are tabulated in Table 3.

**Table 3.** .Primary and Secondary Outcomes at 30 Days.

| Outcome | Immediate Complete Revascularization (N=459) |  | Staged Complete Revascularization (N=458) |  | Hazard Ratio (95% CI) | Risk difference (95% CI) ‡ | P Value |
| --- | --- | --- | --- | --- | --- | --- | --- |
|  | No. events | Percentage † | No. events | Percentage † |  |  |  |
| **Primary outcome** |  |  |  |  |  |  |  |
| All-cause mortality, any myocardial infarction, unplanned ischemia driven revascularization or cerebrovascular event | 8 | 1.8% | 26 | 5.7% | 0.30 (0.13, 0.66)* | 4.0% (1.5, 6.4) | 0.002 |
| **Secondary outcomes** |  |  |  |  |  |  |  |
| Cardiovascular mortality or myocardial infarction | 2 | 0.4% | 15 | 3.3% | 0.13 (0.03, 0.57) | 2.9% (1.1, 4.6) | 0.001 |
| All-cause mortality | 2 | 0.4% | 2 | 0.4% | 1.00 (0.14, 7.07) | 0.0% (-0.9, 0.9) | >0.99 |
| Cardiovascular mortality | 1 | 0.2% | 2 | 0.4% | 0.50 (0.05, 5.49) | 0.2% (-0.5, 1.0) | 0.56 |
| Any myocardial infarction | 1 | 0.2% | 14 | 3.1% | 0.07 (0.01, 0.53) | 2.9% (1.2, 4.5) | <0.001 |
| Unplanned ischemia driven revascularization | 4 | 0.9% | 17 | 3.7% | 0.23 (0.08, 0.68)* | 2.9% (0.9, 4.8) | 0.004 |
| Cerebrovascular event | 2 | 0.4% | 7 | 1.5% | 0.28 (0.06, 1.36) | 1.1% (-0.2, 2.4) | 0.09 |
| Probable or definite stent thrombosis | 2 | 0.4% | 3 | 0.7% | 0.66 (0.11, 3.97) | 0.2% (-0.7, 1.2) | 0.65 |
| Target vessel revascularization | 4 | 0.9% | 17 | 3.7% | 0.23 (0.08, 0.68)* | 2.9% (0.9, 4.8) | 0.004 |
| Target lesion revascularization | 4 | 0.9% | 15 | 3.3% | 0.26 (0.09, 0.79)* | 2.4% (0.6, 4.3) | 0.010 |
| All-cause mortality, myocardial infarction, stroke or major bleeding (BARC 3 or 5) | 6 | 1.3% | 26 | 5.7% | 0.22 (0.09, 0.54)* | 4.4% (2.0, 6.8) | <0.001 |
| Major bleeding (BARC 3 or 5) | 1 | 0.2% | 5 | 1.1% | 0.20 (0.02, 1.70) | 0.9% (-0.2, 1.9) | 0.10 |

BARC=bleeding academic research consortium. \* The cox proportional hazard assumption was not met † Cumulative incidence at 365 days according to the Kaplan-Meier method. ‡ Based on the Kaplan-Meier estimates. A difference in favour of immediate complete revascularization is presented as a positive value. § This P value was tested for superiority of the risk difference.

The cumulative incidence of the primary composite outcome at 1 year follow-up was 7.9% and 10.1% in the patients randomized to ICR and SCR (risk difference 2.2%, 95% confidence interval [CI −1.5 to 6.0, p = 0.24). The incidence of cardiovascular death at 1 year was similar between the two trial arms (1.1% vs. 0.9%, risk difference −0.2%, 95%CI −1.5 to 1.1, p = 0.75). The composite of cardiovascular death and MI occurred in 3.1% and 5.7% of the patients at 1 year, (risk difference 2.7%, 95%CI 0.0 to 5.3, p = 0.052). ICR was associated with a lower incidence of MI (2.0% vs. 5.3%, risk difference 3.3%, 95%CI 0.9 to 5.7, p = 0.008) and unplanned ischemia driven revascularization (4.2% vs. 7.8%, risk difference 3.5%, 95%CI 0.4 to 6.6, p = 0.025) at 1 year. The primary and secondary outcomes at 1 year are tabulated in Table 4.

**Table 4.** Primary and Secondary Outcomes at 1 Year.

| Outcome | Immediate Complete Revascularisation<br>(N=459) |  | Staged Complete Revascularisation<br>(N=458) |  | Hazard Ratio<br>(95% CI) | Risk difference<br>(95% CI) ‡ | P Value |
| --- | --- | --- | --- | --- | --- | --- | --- |
|  | No. events | Percentage † | No. events | Percentage † |  |  |  |
| **Primary outcome** |  |  |  |  |  |  |  |
| All-cause mortality, any myocardial infarction, unplanned ischemia driven revascularization or cerebrovascular event | 36 | 7.9% | 46 | 10.1% | 0.75 (0.48, 1.16)* | 2.2% (-1.5, 6.0) | 0.24 |
| **Secondary outcomes** |  |  |  |  |  |  |  |
| Cardiovascular mortality or myocardial infarction | 14 | 3.1% | 26 | 5.7% | 0.52 (0.27, 1.00)* | 2.7% (0.0, 5.3%) | 0.052 |
| All-cause mortality | 7 | 1.5% | 5 | 1.1% | 1.39 (0.44, 4.38) | -0.4% (-1.9, 1.1) | 0.57 |
| Cardiovascular mortality | 5 | 1.1% | 4 | 0.9% | 1.24 (0.33, 4.62) | -0.2% (-1.5, 1.1) | 0.75 |
| Myocardial infarction | 9 | 2.0% | 24 | 5.3% | 0.36 (0.17, 0.78)* | 3.3% (0.9, 5.7) | 0.008 |
| Unplanned ischemia driven revascularization | 19 | 4.2% | 35 | 7.8% | 0.52 (0.30, 0.91)* | 3.5% (0.4, 6.6) | 0.025 |
| Cerebrovascular event | 7 | 1.6% | 8 | 1.8% | 0.86 (0.31, 2.38)* | 0.2% (-1.5, 1.9) | 0.81 |
| Probable or definite stent thrombosis | 2 | 0.4% | 5 | 1.1% | 0.40 (0.08, 2.05) | 0.7% (-0.5, 1.8) | 0.25 |
| Target vessel revascularization | 16 | 3.6% | 33 | 7.3% | 0.47 (0.26, 0.85)* | 3.8% (0.8, 6.7) | 0.013 |
| Target lesion revascularization | 13 | 2.9% | 30 | 6.7% | 0.42 (0.22, 0.80)* | 3.8% (1.0, 6.6) | 0.007 |
| All-cause mortality, myocardial infarction, stroke or major bleeding (BARC 3 or 5) | 30 | 6.6% | 40 | 8.8% | 0.72 (0.45, 1.15)* | 2.2% (-1.2, 5.7) | 0.21 |
| Major bleeding (BARC 3 or 5) | 8 | 1.8% | 9 | 2.0% | 0.88 (0.34, 2.28) | 0.2% (-1.5, 2.0) | 0.79 |

BARC=bleeding academic research consortium. \* The cox proportional hazard assumption was not met † Cumulative incidence at 365 days according to the Kaplan-Meier method. ‡ Based on the Kaplan-Meier estimates. A difference in favour of immediate complete revascularization is presented as a positive value. § This P value was tested for superiority of the risk difference.

An analysis excluding procedure related MIs occurring during the index or staged procedure was performed due to the possibility of a potential bias caused by the difficulty of diagnosing type 4a MIs during the index event. This analysis consistently showed a significant reduction of MIs in the ICR group (2.0% vs. 4.4%, risk difference 2.4%, 95%CI 0.1 to 4.7, p = 0.039). A total of 13 non procedure related infarctions occurred between the index and staged procedure, of which 10 were type 1, 1 was type 2 and 2 were type 4b MIs. The primary and secondary outcomes at 1 year, excluding type 4a MIs related to the index or staged procedure, are tabulated in Table 5.

**Table 5.** Clinical Outcomes Excluding Index and Staged Procedure Related Myocardial Infarctions

| Outcome | Immediate Complete Revascularization (N=459) |  | Staged Complete Revascularization (N=458) |  | Hazard Ratio (95% CI) | Risk difference (95% CI) ‡ | P Value |
| --- | --- | --- | --- | --- | --- | --- | --- |
|  | No. events | Percentage † | No. events | Percentage † |  |  |  |
| All-cause mortality, myocardial infarction, unplanned ischemia driven revascularization or cerebrovascular event | 36 | 7.9% | 43 | 9.5% | 0.80 (0.52, 1.25)* | 1.6% (-2.1, 5.2) | 0.40 |
| Cardiovascular mortality or myocardial infarction | 14 | 3.1% | 22 | 4.9% | 0.62 (0.32, 1.21)* | 1.8% (-0.8, 4.3) | 0.17 |
| Myocardial infarction | 9 | 2.0% | 20 | 4.4% | 0.44 (0.20, 0.96)* | 2.4% (0.1, 4.7) | 0.039 |
| All-cause mortality, myocardial infarction, stroke or major bleeding (BARC 3 and 5) | 30 | 6.6% | 37 | 8.2% | 0.78 (0.48, 1.26) | 1.6% (-1.8, 5.0) | 0.37 |

BARC=bleeding academic research consortium. \* The cox proportional hazard assumption was not met † Cumulative incidence at 365 days according to the Kaplan-Meier method. ‡ Based on the Kaplan-Meier estimates. A difference in favour of immediate complete revascularization is presented as a positive value. § This P value was tested for superiority of the risk difference.

## Discussion

The current further analysis of the BIOVASC trial, which was prespecified in the trial protocol, suggests a reduction in the incidence of MIs and unplanned ischemia driven revascularizations at 1 year post index PCI when performing ICR in the NSTE-ACS population. The reduction in myocardial infarction associated with an ICR strategy persisted after exclusion of procedure-related events.

In the BIOVASC trial, 44% (N=15) of all first occurring non procedure related MIs in the SCR group, happened between the index and staged procedure. Ten of those MIs were type 1 MI and occurred only in patients that initially presented with a NSTE-ACS at randomization.

Plaque vulnerability of non-culprit lesions might have a role in the occurrence of early spontaneous infarctions in patients with ACS. Several factors could induce plaque instability in the acute phase, such as an enhanced general inflammatory status, oxidative stress, which is an imbalance between the generation of reactive oxygen species and its clearance through the intrinsic antioxidant defense system^20^. Acute MI has been associated with a decrease in antioxidant enzymes^21^, potentially impacting plaque vulnerability in non-culprit lesions. Several studies in ACS and MVD patients^22, 23^ showed the presence of thin-cap fibroatheroma in up to 40% of the analyzed obstructive non-culprit lesions, which is associated with a higher risk of future cardiac events^24^. The non-culprit lesion vulnerability remains yet to be fully evaluated in NSTE-ACS, but a role of diffuse inflammation and plaque instability cannot be excluded in the pathogenesis of the early ischemic events in our population.

Another distinct mechanism that could also explain early ischemic events is the incorrect culprit lesion identification during the index procedure. At variance with STEMI patients in whom the culprit lesion is angiographically evident in the vast majority of the cases, in NSTE-ACS and multivessel disease, culprit lesion assessment can be very challenging^25, 26^. Despite the fact that unclear culprit lesion was an exclusion criteria in the BIOVASC trail, misjudgment of the culprit lesion could have occurred, leading to some acute plaques being left untreated possibly triggering a second early event between the index and staged procedure^27^.

This difference in culprit lesion identification between STEMI and NSTE-ACS patients might also explain the dissimilar progression of the time-to-event curves in this study compared with the COMPLETE trial^19^ in which in the culprit-only revascularization group, events accrued over time in the long-term follow-up.

The SMILE trial showed a significant reduction of the composite of mortality, MI, re-hospitalization for unstable angina, target vessel revascularization and stroke at 1 year when performing ICR instead of SCR in patients presenting with NSTE-ACS and MVD^15^. This effect was driven by a lower risk of target vessel revascularization in the ICR group. In contrast to our study, the time-to-event curves did not diverge early in the follow-up period, but only after 100 days. This discrepancy might be caused by the different study designs. In our study the median time to the staged procedure was 15 days, which is a longer interval than the mean 4.8 days in the SMILE trial, potentially leading to more events in the 30 day timeframe. However, when comparing the results of the SMILE study with ours, the difference in total event rates must also be taken into account. Our study showed a total event rate of 8.9% for the primary composite endpoint, as opposed to 18.4% in the SMILE study driven by a remarkably high rate of target vessel revascularization (15.4% at 1 year follow-up) ^28^.

Similarly to our study, an analysis from the CREDO-Kyoto registry showed significantly lower myocardial infarctions and revascularizations occurring in the ICR group at 30 days post index PCI^29^. At 5 years the study showed no difference in the composite primary outcome or any of its individual components, but both the incidence curves and 30-day results, suggest a similar temporal progression of events compared with our study.

Our data support the adoption of an ICR approach in NSTE-ACS and MVD. In this sub-population of the BIOVASC trial the clinical benefit of ICR was evident in terms of MIs and unplanned ischemia-driven revascularizations regardless of procedure-related events. In addition, similarly to the BIOVASC trial, in the present subanalysis the ICR approach was associated with a reduction in total hospital stay, suggesting possible health economic implications in NSTE-ACS patients^30^.

### Limitations

This is a pre-specified post-hoc analysis of a randomized noninferiority trial. No formal power calculation was performed for this analysis. The use of intracoronary imaging was low, reflecting the current European clinical practice. A higher adoption of imaging might have had an impact on culprit lesion identification providing further insights on the mechanism of early ischemic events.

## Conclusions

In patients presenting with NSTE-ACS and MVD, immediate complete revascularization was safe and associated with a lower cumulative incidence of myocardial infarctions and unplanned ischemia driven myocardial infarction at 1 year post index PCI compared with staged complete revascularization.

## Data Availability

The data from the BIOVASC trial can't be shared owing to the contract stipulated with the grant giver and also with internal regulations at the Erasmus Medical Center, but researchers interested in collaboration should contact the corresponding author.

## List of Abbreviations

ACS: acute coronary syndrome
CI: confidence interval
ICR: immediate complete revascularization
MVD: multivessel disease
MI: myocardial infarction
NSTE-ACS: non-ST-segment elevation acute coronary syndrome
NSTEMI: non-ST-segment elevation myocardial infarction
PCI: percutaneous coronary intervention
PH: proportional hazards
SCR: staged complete revascularization
STEMI: ST-segment elevation myocardial infarction
UA: unstable angina

